# Uncertainty Quantification in Cost-effectiveness Analysis for Stochastic-based Infectious Disease Models: Insights from Surveillance on Lymphatic Filariasis

**DOI:** 10.1101/2024.07.31.24311315

**Authors:** Mary Chriselda Antony Oliver, Matthew Graham, Ioanna Manolopoulou, Graham F. Medley, Lorenzo Pellis, Koen B Pouwels, Matthew Thorpe, T. Deirdre Hollingsworth

**Affiliations:** University of Cambridge, Department of Applied Mathematics and Theoretical Physics, Wilberforce Road, Cambridge, CB3 0BN, UK; University College London, Department of Statistical Science, Gower Street, London, WC1E 6BT, UK; The Alan Turing Institute, London, NW1 2DB, UK; London School of Hygiene and Tropical Medicine, Centre for Mathematical Modelling of Infectious Disease and Department of Global Health and Development, Keppel Street, London, WC1E 7HT, UK; University of Manchester, Department of Mathematics, Oxford Road, Manchester, M13 9PL, UK; University of Oxford, Health Economics Research Centre, Nuffield Department of Population Health, Oxford, OX3 7LF, UK; University of Warwick, Department of Statistics, Coventry, CV4 7AL, UK; University of Oxford, Big Data Institute, Li Ka Shing Centre for Health Information and Discovery, Oxford, OX3 7LF, UK

**Keywords:** Lymphatic filariasis, mathematical modelling, stopping threshold, cost-effective analysis, optimal transport

## Abstract

Cost-effectiveness analyses (CEA) typically involve comparing effectiveness and costs of one or more interventions compared to standard of care, to determine which intervention should be optimally implemented to maximise population health within the constraints of the healthcare budget. Traditionally, cost-effectiveness evaluations are expressed using incremental cost-effectiveness ratios (ICERs), which are compared with a fixed willingness-to-pay (WTP) threshold. Due to the existing uncertainty in costs for interventions and the overall burden of disease, particularly with regard to diseases in populations that are difficult to study, it becomes important to consider uncertainty quantification whilst estimating ICERs.

To tackle the challenges of uncertainty quantification in CEA, we propose an alternative paradigm utilizing the Linear Wasserstein framework combined with Linear Discriminant Analysis (LDA) using a demonstrative example of lymphatic filariasis (LF). This approach uses geometric embeddings of the overall costs for treatment and surveillance, disability-adjusted lifeyears (DALYs) averted for morbidity by quantifying the burden of disease due to the years lived with disability, and probabilities of local elimination over a time-horizon of 20 years to evaluate the cost-effectiveness of lowering the stopping thresholds for post-surveillance determination of LF elimination as a public health problem. Our findings suggest that reducing the stopping threshold from <1% to <0.5% microfilaria (mf) prevalence for adults aged 20 years and above, under various treatment coverages and baseline prevalences, is cost-effective. When validated on 20% of test data, for 65% treatment coverage, a government expenditure of WTP ranging from $500 to $3,000 per 1% increase in local elimination probability justifies the switch to the lower threshold as cost-effective.

Stochastic model simulations often lead to parameter and structural uncertainty in CEA. Uncertainty may impact the decisions taken, and this study underscores the necessity of better uncertainty quantification techniques within CEA for making informed decisions.

## 1. Introduction

### 1.1. Health Economics Motivation

Global health systems face enormous challenges as a result of the rising demand for healthcare services and the finite resources available to them. While other factors such as equity may play a role, a common aim for governments is to maximise overall population health within the constraints of the available healthcare budget. Planning, managing, and assessing health systems heavily relies on economic factors. The best use of limited resources is guided by health economic analyses, which provide cohesive techniques for evaluating the cost-effectiveness of health interventions.

The economic evaluation of health interventions is normally based on the outcome and cost of the interventions. Depending on the choice of how the outcome and intervention is evaluated, one of the main methodologies used is the cost-effectiveness analysis (CEA). This is an economic evaluation technique in which two or more health interventions are compared in terms of incremental costs and incremental effects compared to standard of care, with the cost-effectiveness expressed using the incremental cost-effectiveness ratio (ICER) which is a measure dividing the incremental costs by the incremental effects. Most countries that regularly use CEA to guide policy decisions around the implementation and reimbursement of interventions specify in their health-economic guidelines that cost-utility analyses should be used, where the denominator of the ICER is expressed in quality-adjusted life-years (QALYs) or disability-adjusted life-years (DALYs). The latter is more frequently used in low- and middle-income countries (LMICs). Given the focus on LMICs in this paper, DALYs will be used from here onwards.

The interpretation of the ICER depends on where it lies on the cost-effectiveness plane (refer Figure 1 in [20]). If the new intervention is more effective and saves money compared to standard of care (South East quadrant), or the new intervention is less effective and more costly compared to standard of care (North West quadrant), the ICER is negative and interpretation is simple. In the former case the new intervention should clearly be adopted from a cost-effectiveness point of view, whereas in the latter case the intervention is clearly worse and should not be adopted.

**Figure 1:**
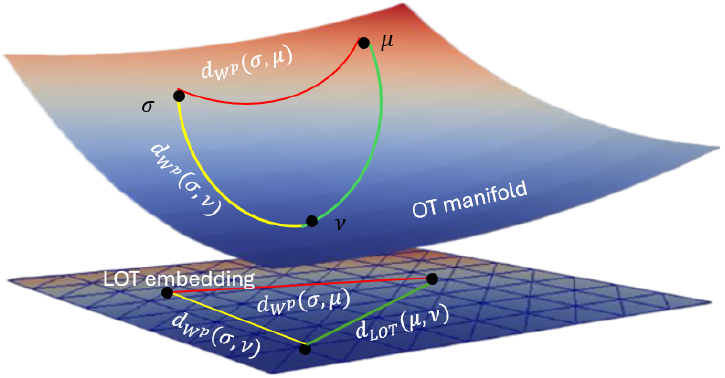
The Linear *p*-Wasserstein framework embeds measures in the tangent space of a fixed reference σ. As a consequence, the Euclidean distance between the non-negative measures *µ* and *ν* is an approximation for the 2-Wasserstein distance 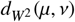. This figure is computed using ParaView.

To determine whether an intervention likely improves overall population health within the healthcare budget constraints, the ICER can be compared with a cost-effectiveness threshold in situations where the intervention is more costly and more effective (North East quadrant) or less costly and less effective (South West quadrant). Assuming the decision-maker indeed wants to maximise overall population health, symmetrical threshold should be applied for both quadrants, whereby interventions that are more effective and more costly should remain below the threshold and interventions that are less effective and less costly should remain above the threshold.

If a new intervention costs more per DALY avoided than the healthcare it displaces, health opportunity costs exceed health benefits, and implementing the new intervention would be expected to lead to an overall reduction in population health measured in DALYs. Theoretically, the cost-effectiveness threshold [12; 48; 35] should reflect the point at which this occurs. Thus, given the available budget, interventions that are more costly and more effective with an ICER below the threshold are expected to improve overall population health, while similar interventions with an ICER above the threshold are expected to worsen overall population health.

Characterizing uncertainty is crucial in CEA, particularly when evaluating the need for additional evidence. Value of Information (VoI) analysis enhances CEA by quantifying the benefit of reducing uncertainty in decision-making. In health decision-analytic models, VoI assesses the potential benefit obtaining additional data aimed at reducing uncertainty in key parameters influencing decision uncertainty. Two key uncertainties are model input values and model structure, whereby VoI analyses in the literature typically only focus on parameter uncertainty and completely ignore model structure uncertainty. These models are typically law-driven due to a lack of long-term data. To quantify input uncertainty, a probability distribution for true input values is propagated through the model using Monte Carlo sampling, known as probabilistic sensitivity analysis (PSA) [15; 43]. However, PSA only addresses input uncertainty, not structural uncertainty, which is harder to quantify and requires judgments about the model’s real-life representation.

Despite its potential, VoI analysis [56] is constrained by structural uncertainties, which are rarely quantified in model-based analyses. Not quantifying structural uncertainty implies that the model is a perfect representation of real-world processes and relationships. While VoI analysis for structural uncertainty using model selection and model averaging has been explored previously [44; 6], methods in this area are still underdeveloped. Addressing these limitations is essential to fully leverage VoI analysis in making informed and effective healthcare policy decisions.

### 1.2 Theoretical Background on Lymphatic Filariasis

Lymphatic filariasis (LF), a debilitating neglected tropical disease caused by parasitic worms transmitted through mosquitoes, affects about 882 million people across 44 countries [54]. In 2000, the World Health Organization (WHO) launched the Global Program to Eliminate Lymphatic Filariasis (GPELF), aiming to eradicate LF as a public health problem (EPHP) in 73 endemic nations by 2020 [53]. By 2021, 19 countries, including Bangladesh and Lao People’s Democratic Republic, were validated as having achieved EPHP, with 11 others under surveillance after halting large-scale treatment [53; 54].

The primary intervention involves annual mass drug administration (MDA) for at least five years in affected areas, employing drug combinations such as diethylcarbamazine (DEC) + albendazole (DA) or albendazole + ivermectin (IA) [54]. Some areas also utilize a triple combination ivermectin + DEC + albendazole (IDA) [28; 24]. To assess MDA impact and determine if infection levels have dropped below stopping thresholds, WHO recommends epidemiological monitoring surveys and transmission assessment surveys (TAS). The TAS uses the samples of blood smears, typically surveying children aged 5 years and above for microfilariae (mf) prevalence [54]. Current MDA guidelines advise a minimum of 5 rounds of treatment before a pre-TAS is used to determine whether a first full TAS should be conducted, known as TAS-1. MDA can be stopped if TAS-1 is passed. Two subsequent surveys must also be passed before EPHP can be validated, TAS-2 and TAS-3, each within 2–3 years of the previous assessment.

However, focusing solely on children may underestimate mf prevalence, potentially missing ongoing transmission due to higher mf prevalence in adults. This paper proposes to improve the sensitivity of TAS to evaluate mf prevalence in adults, targeting <0.5% mf prevalence. This involves randomly sampling approximately 30 sites with 40-60 adults per site to replicate the characteristics of an evaluation unit (EU). Achieving and sustaining WHO goals necessitates effective surveillance, identifying new cases post-EPHP target attainment. Intensive surveillance thresholds (<2% antigenamia (Ag), <1% mf) may still be inadequate, especially in areas with *Culex* transmission vectors [3; 16]. Mathematical and biological theories [1] propose a transmission breakpoint influenced by local transmission conditions and biological factors, in helminth infections such as LF which depend on sexual reproduction of the parasites, where low worm burdens diminish onward transmission, potentially leading to disease extinction in deterministic scenarios. Studies, have suggested that the breakpoint might be substantially lower than 1% mf prevalence [19; 34]. Stochastic extinction can still occur above this breakpoint but with a lower probability [16]. If MDA are halted after reaching the breakpoint, the low-level remaining transmission will diminish gradually taking a longer time for LF extinction.

In this work, we aim to provide the first detailed model simulations of reducing the TAS stopping threshold in LF from <1% to <0.5% mf prevalence in a sample of adults aged 20 years and above. This facilitates the understanding of the different trade-offs between additional rounds of MDA treatment and rebounds that apply to the design of surveillance strategies. In this context, modelling can help us to understand how adjusting the threshold used in TAS impacts decisions about the stop of interventions and at what cost. For many settings, a reduction in the threshold increases the probability of elimination, decreases the number of treatment rounds required, and reduces costs. Importantly, however, in certain circumstances (e.g., when coverage is lower), lower thresholds can imply an increase in the number of rounds of treatment required to reach that threshold (with increased costs) but help mitigate chronic conditions (such as lymphoedema and hydrocele) and result in longer sustained elimination with fewer future rebounds.

To investigate the issues outlined above, here we use mathematical models of the transmission dynamics of LF as a case study to assess the potential implications of modifying the threshold for TAS. The paper addresses a key question: What are the potential trade-offs encountered in uncertainty quantification of cost-effectiveness analysis on lowering the stopping threshold for TAS in adults aged 20 years and above from an economic, epidemiological and mathematical perspective? In this paper, we restrict to lowering the stopping threshold from <1% mf prevalence to <0.5% mf prevalence for a sample of adults motivated by the work in [3] and [16]. Importantly, we focus on areas with *Culex* mosquitoes as the major transmission vector using IA drug combinations for potential comparisons.

### 1.3. Contributions

This study investigates the following three specific sub-questions outlined below which highlight the key contributions of our work:

1. What is the interplay between the dynamics of infection on DALY burden and elimination? In Section 5 we show the monotonic behaviour of the DALY burden and probability of elimination for different stopping thresholds, baseline mf prevalences and MDA coverages.
2. What are the dynamics of the costs both pre and post-MDA surveillance when the stopping threshold is lowered? In Section 5 we explain the tradeoff illustrated in the observed non-monotonic behaviour of the costs for different stopping thresholds, baseline prevalences and MDA coverages.
3. If lower stopping thresholds are required for elimination of transmission, then are we realistically able to measure them using current tools? In order to circumvent issues related to the ICERs to address this question using the CEA framework in Section 2, we instead adopt a linear formulation of Expected Incremental Net-Monetary Benefit (EINMB) metric for fixed country-level WTP values as recommended by the several studies [38; 52] for DALYs averted and approximate the range of WTP for probability of elimination (due to lack of data) in order to align with the goals of GPLEF. We also extend the analysis to quantify uncertainty with every additional sample size using Value of Information Analysis (VoI) with the help of Expected Value of Sample Size (EVSI) metric for the optimum WTP values per DALY averted and unit increase in the probability of elimination for different stopping thresholds and baseline prevalences. Finally, we propose an alternate paradigm, the Linear Wasserstein Framework in Section 3 that might help us resolve some of the proposed limitations, particularly around structural uncertainty of the CEA framework.

Addressing these questions will help to assess whether lower thresholds have the potential to assist programmes in achieving LF local elimination goals and how such decisions impact programme costs aligning with the GPELF objectives.

### 1.4. Outline of the paper

We begin in Section 2 by summarizing the theoretical framework for CEA. In addition, we prescribe an alternative paradigm that circumvents structural uncertainties of CEA using the Linear Wasserstein framework in Section 3. The numerical implementation is then described in Section 4 and tested in Section 5. Finally, we will discuss our findings and present our conclusions in Section 6. For the reader’s convenience we have provided a list of key terminologies used in the manuscript in Table C.15 (see Appendix C for more details).

## 2 Summary on Cost Effective Analysis (CEA) Framework

In this section, we extend the classic net-benefit framework to include resource implications alongside aligning with GPLEF goals by incorporating elimination probabilities [2]. Additionally, we incorporate Value of Information (VoI) analysis which directly addresses the potential implications of current uncertainty, not only in terms of the likelihood of modifying the current decision in light of new and more definitive evidence, but also in terms of the opportunity cost of the incorrect decision.

### 2.1. Notation and Basic Concepts

Health economic decision making aims to determine the optimal intervention considering costs and health impacts of various clinical effectiveness. A key metric is the Incremental Cost-Effectiveness Ratio (ICER), defined as the ratio of the difference in costs (Δ*C*) to the difference in health impacts (Δ*E*) between two interventions:

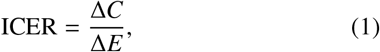

Here, we use a cost-utility analysis, where health impacts (ΔE) are expressed in disability-adjusted life years (ΔDALYs) averted by quantifying overall disease burden due to morbidity and mortality. In the current analyses, we only include effects on morbidity (e.g., lymphoedema, hydrocele) as we assumed the intervention has no impact on mortality. For our analysis, we will use both DALYs averted and the probability of elimination. A strategy is considered cost-effective if the ICER does not exceed the health planner’s WTP per DALY averted (WTP_DALY_),

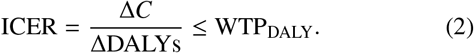

The net-benefit framework circumvents issues with ICERs by not having to deal with extended dominance (when one intervention is less cost-effective than a combination of two or more interventions) by transforming the ICER into a linear additive form, known as the net-monetary benefit (NMB).

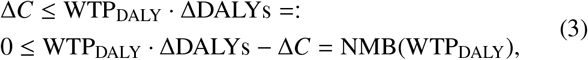

By using NMB, which relies on single monetary values rather than ratios, the framework simplifies the evaluation of multiple interventions, regardless of which quadrant of the cost-effectiveness plane the ICER lies in. Given a Monte Carlo sample of *N* iterates of the costs and DALYs averted (denoted by the parameter set *θ*), a strategy is preferred over the comparator if the expected NMB exceeds zero:

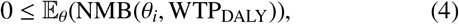

where *θ* is the parameter vector and *i* = {1, 2, ⋯, *N*} is the iterations per parameter to denote the samples drawn from the joint distribution *p*(θ). Extending this framework to multistrategy decision analysis between *J* strategies, the preferred strategy is the one that maximizes the 𝔼_θ_(NMB(θ_*i*_, WTP_DALY_)):

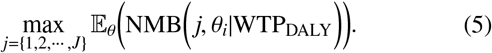

We also include benefits related to the probability of elimination of LF, aligning with GPELF goals using WTP per unit increase in probability of elimination (WTP_Elimination_). The NMB is reformulated as:

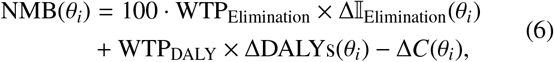

Here, Δ𝕀_Elimination_(θ_*i*_) is 1 if only one strategy achieves elimination, and 0 otherwise such that 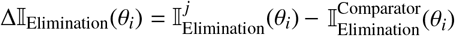. Analogous to the traditional NMB, the strategy that ought to be implemented is indicated by,

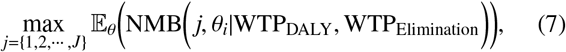

Simultaneously, the framework allows for a probabilistic interpretation of cost effectiveness by conditioning on WTP_DALY_, WTP_Elimination_ as follows,

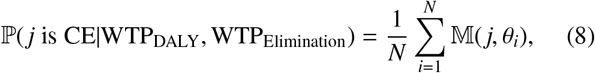

where

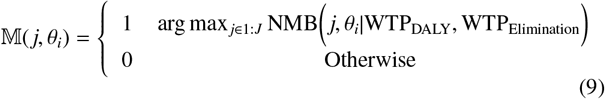

The framework therefore presents a measure of uncertainty that the strategy with the highest expected NMB is optimal over all other strategies, given by the proportion of samples where the strategy has the highest NMB of all strategies.

In general terms, the Expected Value of Perfect Information (EVPI) [8] is the difference between the expected value of a decision made with perfect information and the expected value of a decision made with current knowledge. It represents the maximum amount a decision-maker would be willing to pay for perfect information to avoid the potential losses associated with uncertainty. EVPI is defined as,

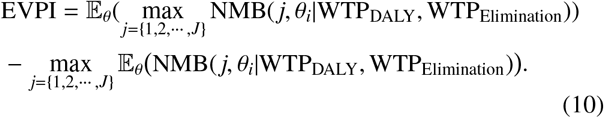

Here, the vector of parameters can be split in two components *θ* = (*ϕ, ψ*), where *ϕ* is the subvector of parameters of interest (i.e., those that could be investigated further) and *ψ* are the remaining “nuisance” parameters:

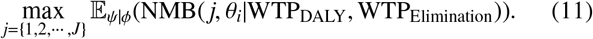

which is the value of learning *ϕ* with no uncertainty. Of course, we will never be in the position to completely eliminate the uncertainty on *ϕ*, so we then average over its current probability distribution while also subtracting the value of the current optimal decision to calculate the Expected Value of Partial Perfect Information (EVPPI) [8; 43; 22]. The economic value of eliminating all uncertainty about *ϕ* (assuming risk neutrality) is equal to the EVPPI which is given by:

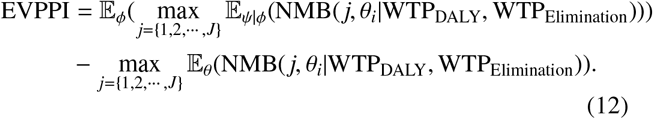

Expected Value of Sample Information (EVSI) [23] measures the value of collecting additional data *X* to inform *ϕ*, assuming *X* directly updates *ϕ* and is independent of ψ|*ϕ*. EVSI is bounded above by EVPPI. If data *X* were observed as *x*, it would update *ϕ*’s distribution *p*(*ϕ*|*x*), impacting the net benefit distribution for each treatment. EVSI is the average value over all possible data sets:

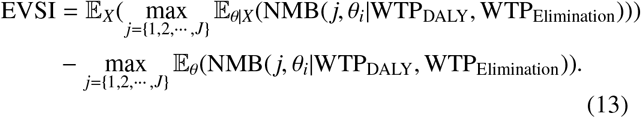

In this paper, to estimate EVSI computationally we follow the efficient nested Monte Carlo method based on “moment matching” by [23]. The method improves computational efficiency by reducing the nested Monte Carlo error, using the moment matching technique to approximate the distribution of the posterior samples more accurately (see for more details, Algorithm 1 in Appendix A). Although, several other approaches [29] such as the Importance Sampling (IS), Regression techniques, Gaussian approximation method and Integrated Nested Laplace Analysis (INLA) [22] exist, we rely on moment matching for its ability to estimate EVSI for multiple alternative sample sizes with a fixed additional computational cost. We also note that Integrated Nested Laplace Analysis (INLA) is highly efficient for performing Bayesian inference, especially in latent Gaussian models by treating the PSA simulations as a ‘spatial problem’ and projecting from higher to lower dimension to evaluate EVPPI by dimensionality reduction.

### 2.2. Fundamental Issues using EVSI as a metric

There are several challenges that arise when using the EVSI metric:

1. Assumptions on Distributions: Implementing EVSI using the moment-matching method (see Algorithm 1 in Appendix A) involves approximating the distribution of the sample information using moments (mean, variance, etc.). This can introduce errors, especially if the true distribution of the sample information is not well-approximated by the moments.
2. Dependence on Prior Information: The quality of EVSI estimates using moment matching depends heavily on the prior information available. Poor or inaccurate priors can lead to misleading EVSI estimates.
3. Implementation Challenges: Moment matching is accurate and efficient when the health economic model has a low computation time but becomes more unfeasible as the model runtime increases and inaccurate when the sample size is less than 10.
4. Requires Accurate EVPPI Estimation: Moment matching is more accurate for studies that will have significant impact on the underlying uncertainty in the decision-analytic model, i.e., the EVPPI of *ϕ* needs to be high compared to the value of reducing all model uncertainty (i.e., EVPI), ideally greater than 40% [23].

While moment matching can be a useful tool for approximating EVSI, these limitations must be carefully considered and addressed to ensure accurate and reliable health economic decision-making.

## 3. Linear Wasserstein Framework

This section introduces an alternative metric to EVSI for determining the cost-effectiveness of intervention strategies using the Linear Wasserstein Framework (also called “Linear Optimal Transport” (LOT)) which was originally formulated in [55]. Wasserstein-like distances are metrics on probability measures. They can be motivated from a geometric point-of-view as they pay a cost based on rearrangement of mass. This means the distance is assigned (loosely speaking) based on translations. The main obstacle concerning Wasserstein distances is the computational cost and a lack of off-the-shelf data analysis tools. This is where the linearisation of optimal transport distances plays an important role. The linearisation defines a map *P* : 𝒫(*X*) → ℝ^*k*^ (for some *k*) such that the Wasserstein distance in 𝒫(*X*) is approximately the Euclidean distance in ℝ^*k*^. In the Euclidean space we can easily apply several standard data analysis tools such as dimensionality reduction, classification and modelling.

### 3.1. Notations

Let *X, Y* ⊆ ℝ^*d*^ with *µ* ∈ 𝒫(*X*) and *ν* ∈ 𝒫(*Y*). Given *µ* ∈ 𝒫(*X*) and a transport map *T* : *X* → *Y* we can define the pushforward of *µ* by *T* as follows.

#### Definition 1

*Let µ* ∈ 𝒫(*X*) *and T* : *X* → *Y be a measurable map, the pushforward of µ by T, denoted as T*_#_*µ is the measure ν defined by*,

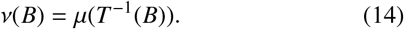

*for all measurable set B* ⊆ *Y*.

### 3.2. Optimal Transport Formulations

Let us consider *T* : *X* → *Y* to be a Borel measurable function such that *T*_#_*µ* = *ν*. The *Monge formulation* [11] would be to find the transport map *T*, given the probability measures *µ,ν*, minimising the objective function in 𝕄(*µ, ν*), where

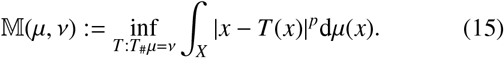

We call any *T* which satisfies *T*_#_*µ* = *ν* a transport map and the minimizer of the optimisation problem in Eq. (15) as the optimal transport map *T*\*. It is often difficult to handle this non-convex optimisation problem in Eq. (15) due to its non-linearity in *T*.

We define the set Π(*µ, ν*) of couplings between measures *µ* and *ν* to be the set of probability measures on the product space 𝒫(*X* × *Y*) whose first marginal is *µ* and the second marginal is *ν*. For any transport map *T* : *X* → *Y*, there exists an associated transport plan *π* such that,

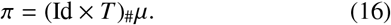

where Id denotes the identity map. We recall that if *P*^*X*^ : *X* × *Y* → *X* and *P*^*Y*^ : *X* × *Y* → *Y* are the canonical projections, then the marginals are 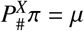 and 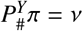.

The *Kantorovich formulation* [11] would be to minimise the objective function 𝕂(*µ, ν*), given the probability measures *µ, ν*, where

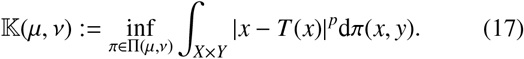

The minimizer of Eq. (17) is the optimal transport plan *π*^∗^. In this sense, the *Kantorovich formulation* in Eq. (17) can viewed as a relaxation of the *Monge formulation*. The difficulty in proving the existence of maps that satisfy the constraint *T*_#_*µ* = *ν* leads to mass splitting during transportation, especially in cases involving discrete measures where such transport maps may not be feasible. This modified formulation in Eq. (17) now describes the amount of mass *π*(*x, y*) that can be transported from *x* to different positions at *y*.

### 3.3. Wasserstein Distances

We denote the space of probability measures on *X* that have finite *p*^*th*^ moment, as follows:

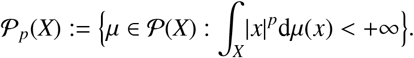

So, when *X* is bounded 𝒫_*p*_(*X*) = 𝒫(*X*). This allows us to define the *p* − Wasserstein distance [11], which is the minimum transportation cost between *µ* and *ν*, as

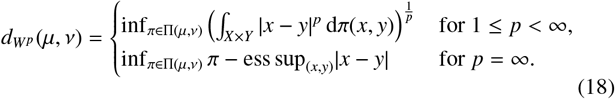

The *d*_*W*_*p* distances are advantageous for Lagrangian modeling due to their simplicity, metric properties (like symmetry), existence of geodesics, Riemannian structure and theoretical benefits like existence of optimal transport maps and plans. However, they require the inputs to be probability measures, are computationally expensive, and there is a lack off-the-shelf data analysis tools. We therefore opt for the Linear *p*-Wasserstein Framework.

### 3.4. Linear Wasserstein Framework

The Linear p-Wassertein framework introduced by Wang in [55], illustrated in Figure 1, has several applications in biomedical imaging, analysis of 2-D point cloud data [5; 36], telescopic and facial expressions [32; 31]. The term “linear” refers to the (Euclidean) vector space structure that one gains after approximation. The method linearizes the Wasserstein distance by computing the projection to tangent space at a fixed reference.

To discuss the Linear p−Wasserstein framework in the continuous setting we consider a domain *X* ∈ ℝ^*d*^ that is a bounded, convex and closed subset of ℝ^*d*^ with a non-empty interior, alongside the probability measures *µ*_*i*_ ∈ 𝒫(*X*) ∀ *i* ∈ {1, 2, ⋯, *N*} and a fixed reference σ ∈ 𝒫(*X*). The optimal transport map 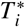 between σ and *µ*_*i*_ satisfies,

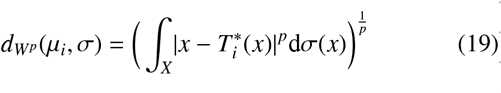

Where 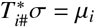. This provides the basis to formally introduce the linear Wasserstein distance for two measures say *µ*_1_ and *µ*_2_.

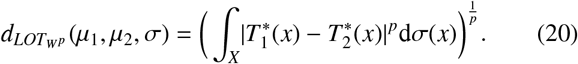

This enables us to compute the linear embeddings in the form of projections *P* : 𝒫(*X*) → *T*_σ_𝒫(*X*) which are the velocity maps from the manifold to the tangent space. This can be expressed as,

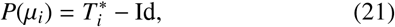

Equivalently, relating the Linear Wasserstein distance 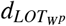 to *p*-Wasserstein distance *d*_*W*_*p* in equation (19) we can rewrite as follows,

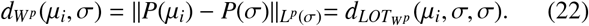

*Remark* 2. This implies that the maps *P*(*µ*_*i*_) form the linear embeddings in the form of projections from the p-Wasserstein space to *L*^2^ (Euclidean) space, thereby preserving the optimal transport distance between *µ*_*i*_ and σ. It is assumed that *d*_LOT_W*p* (*µ*_1_, *µ*_2_, σ) ≈ *d*_W_*p* (*µ*_1_, *µ*_2_) and the approximation depends on the curvature of the Wasserstein space and in general the linear Wasserstein distance is not equivalent (in terms of metric equivalence) to the Wasserstein space. However, when there is some special structure, such as when the measures are all translations or shearings then one gets established bounds [10] like 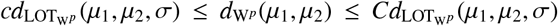 where *c, C* are some positive constants.

### 3.5. Advantages of using the Linear Wasserstein Framework

The linear optimal transport (LOT) framework offers an alternative approach to the calculation of the Expected Value of Sample Information (EVSI), addressing several limitations associated with the moment matching method as outlined below:

1. Approximation Accuracy: Unlike moment matching, which approximates the distribution using moments, LOT can directly handle the full distribution of the data by using the probability measures. This leads to more accurate representations of the underlying distributions, reducing approximation errors. According to ISPOR’s recommendations [39], uncertainty in parameter input values should be characterized using probability distributions. Additionally, any dependencies between parameters should be represented by a joint, correlated probability distribution. We have modeled these inputs as a point cloud (discrete set of points in space).
2. Handling High Dimensions: The LOT framework provides linear projections that help reducing the dimensionality from the ambient to the tangent space after applying off-the-shelf data analysis tools.
3. Distributional Assumptions: LOT does not rely on the assumption that the distribution can be adequately described by expectation and variance. It considers the entire distribution suitable for handling non-linearities and interactions inherent in the data, thus accommodating higherorder moments more naturally by preserving salient properties of the data and accounting structural uncertainty.
4. Independence from Prior Information: The LOT approach minimizes this dependency by utilizing an empirical distribution derived from observed data. This empirical focus means that the approach is less susceptible to the biases introduced by incorrect prior assumptions.

By directly addressing the distribution of sample information and leveraging efficient optimization techniques, the LOT framework provides a robust and scalable uncertainty quantification tool to calculate cost-effectiveness analysis. This circumvents many of the limitations associated with moment matching, leading to more accurate and reliable decision-making.

## 4. Methods

We utilize the stochastic TRANSFIL model [25] with parameters previously estimated [26; 45] to represent transmission by Culicine mosquitoes (for more details on the parameters refer Table C.14 in Appendix C). The model simulates the health impacts of lymphatic filariasis (LF) and incorporates mass drug administration (MDA) effects, based on simulated target coverage, systematic nonadherence, and drug efficacy [18]. We excluded other interventions such as vector control for this study. We modeled closed populations of 100,000-500,000 people, reflecting EU sizes in standard TAS surveys per WHO guidelines [54]. The detection parameters were fitted using Bayesian MCMC to data from Malindi, Kenya, Colombo, Gampaha and Sri Lanka [26]. MDAs were simulated at 65% and 80% coverage. Systematic non-adherence was included by calculating individual treatment probabilities based on coverage and between-round correlation, parameterized with data from Leogane, Haiti, and Egypt [18].

The model also simulates health impacts of lymphoedema, hydrocele, and acute adenolymphangitis (ADL) using published methods. Morbidity due to lymphoedema and hydrocele was modeled with data from India [13]. The model assumes morbidity occurs after accruing a certain cumulative worm burden. ADL incidence was estimated twice per year in 70% of hydrocele patients and four times annually in 95% of lymphoedema patients [14]. Prevalence was converted using published disability weights [21]. Side-effects of MDA were not considered, despite reports of 13% feeling unwell post-MDA, as these effects were deemed minor [59]. Mental illness was also excluded due to lack of accurate data, despite its recognized burden in LF [49; 30].

For WHO-prescribed starting and stopping decisions [54], we considered TAS surveys from 30 sites per EU. Baseline prevalences were sampled from a normal distribution with means of 5-10%, 10-20%, or 20-30%. In each site, we sampled 40-60 adults aged 20 years to evaluate TAS. If mf-positive adults were below the stopping threshold MDA was halted until the next survey; otherwise, it continued. We iterated this algorithm 1,000 times and reported mean baseline prevalences. Cost simulations considered TAS surveys ($12,494.75 [7]) and MDA rounds ($7,640.92 [47]) over a 20-year horizon, with discounting included. We note that for MDA restarts, the costs of the MDA and TAS are doubled.

For cost-effectiveness analysis, using the Expected Incremental Net Monetary Benefit (EINMB) metric, we used <1% mf prevalence in children (aged 5 years and above) as the comparator. We simulated transmission dynamics and morbidity associated with LF, including DALY burden for 30 sites and a TAS-like survey across those sites. We investigated different MDA coverages (65% and 80%) and different baseline LF prevalences. We evaluated WTP_DALY_ for DALYs averted, reflecting opportunity costs and adjusted for purchasing power parity [51] using $500 (Ghana), $2,500 (Congo) and $5,000 (Southern Africa) based on the provided country-specific percentage of GDP per capita estimate that underlies the DALY-4 estimation method by multiplying the total per individual DALY value times a specific proportion of the GDP per capita [35] for LMIC and WTP_Elimination_ per unit increase in local elimination ranging from $0-$10,000 [2]. We evaluated different stopping thresholds for Culicine transmitters using a model-based transmission dynamics, health, and economic impacts. To address our key questions, we examined:

1. Probability of elimination [46], i.e., the probability of achieving local elimination within 20 years post-MDA if mf prevalence in a sample of <1,700 adults aged ≥ 20 years was below the stopping threshold.
2. Health impact evaluation through DALYs averted for morbidity by quantifying the overall disease burden due to lymphoedema, hydrocele and ADL by the years lived with disability. In this context, we assume that the years of life lost due to premature death is zero as death due to LF is rare.
3. Costs due to MDA rounds and TAS surveys.
4. Computing the cost-effectiveness of lowering the stopping threshold to <0.5% in adults with the help of the EINMB metric (see Section 2). We note that for this metric we use the fixed country-specific WTP_DALY_ and vary across an approximate range of WTP_Elimination_.
5. Evaluating the uncertainty in parameters (total costs, DALYs averted and/or probability of elimination) using Value of Information (VoI) methodology, with EVSI metric (see Section 2) implemented using moment matching method (see Algorithm 1 in Appendix A).
6. Evaluating the uncertainty in parameters (total costs, DALYs averted and/or probability of elimination) using the Linear Wasserstein Framework in conjunction with LDA (see Section 3). The total costs due to MDA rounds and TAS surveys, along with DALYs averted and probability of elimination, are represented as probability measures on point cloud data (discrete set of data points in space). Let *µ*_*i*_ denote the empirical measure associated with the *i*-th point cloud, defined as: 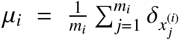 where *x* _*j*_ ∈ ℝ^*d*^ and *m*_*i*_ denotes total data points in each *µ*_*i*_. We sample the point clouds so that each point cloud has the same fixed number *m* = 1000 data points, i.e. 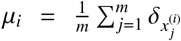 (up to relabelling of the 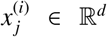) for *i* = {1, 2, ⋯, *N*} and *j* = {1, 2, ⋯, *m*}. Following this, we compute the projections *P*(*µ*_*i*_) as defined in Eq. (21) for each measure *µ*_*i*_. Using these projections, firstly we apply PCA to obtain the eigenvectors that accounts for principle variations of the distributions. We then use these projected eigenvectors from PCA as feature vectors (denoted as 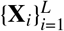) along with three classes of baseline prevalences (5-10%,10-20%,20-30%) as labels (denoted as 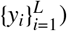 where *L* is the size of training dataset used as inputs for LDA [58] to classify the <0.5% mf threshold for different baseline prevalences relative to the reference. We note that we train the LDA on 80% of the feature vector and labels (iteratively) and make predictions on the remaining 20% of the unseen feature vector and labels. In this study, we use the fixed WTP_DALY_ as outlined above and estimate the range for WTP_Elimination_ (see Algorithm 2 in Appendix A for more details)

## 5. Results

The impact of MDA on the interruption of LF transmission and reduction of the disease burden using DALYs is dependent on the threshold criteria defined for passing the TAS, as illustrated in the example of a setting with a baseline prevalence of 5-10% and 80% MDA coverage of a single population size of 1000 for 10 simulations (Figure 2).

**Figure 2:**
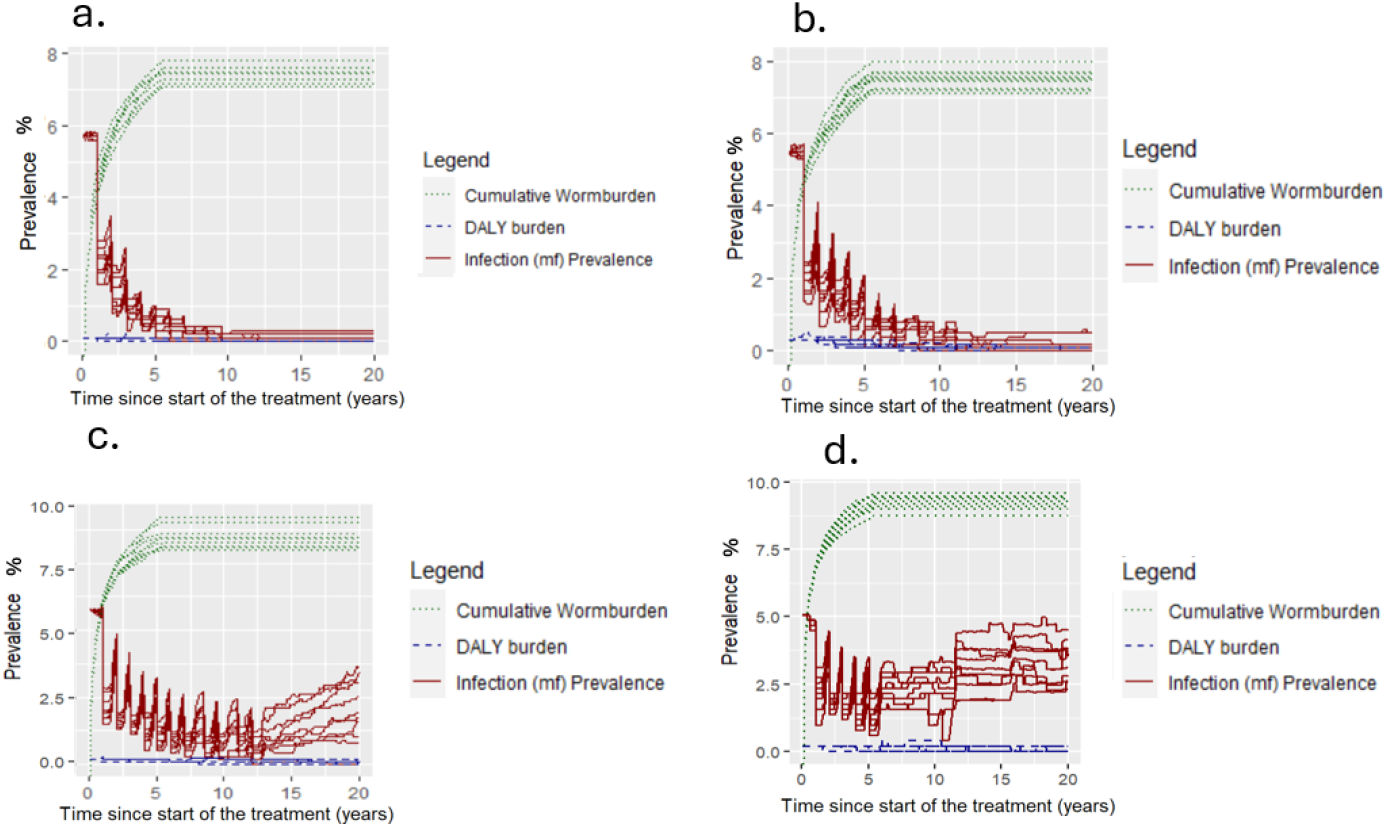
Simplified timeline plots for a single population of size 1000 for 10 simulations illustrating the model-predicted temporal trends in mf prevalence (solid red lines), DALY burden are computed as the morbidity prevalence of lymphoedema, hydrocele and acute adenolymphangitis (dashed blue lines) times the disability weights [21] and cumulative wormburden (dotted green lines) for 5-10% mf prevalence using (a) <0.5%, (b) <1%, (c) <2% and (d) <5% as the stopping threshold criteria for TAS with 80% MDA coverage for a sample of adults.

In Figure 3 (circles) replicating the characteristics of an EU, we find that the probability for local elimination at 5-10% baseline prevalence with 80% MDA coverage and a threshold of <0.5% mf prevalence was 89.2% (≥5 years), 91.8% (≥20 years), and 90.72% (entire eligible population). For a threshold of <1% mf prevalence, it was 80.05%, 83.8%, and 81.76%, respectively. Lowering the threshold increases the probability for local elimination across different prevalences, coverages, and age-groups. These trends follow across different baseline prevalences, MDA coverages and treatment strategies (refer B.1, B.2, and B.3 in Appendix B).

**Figure 3:**
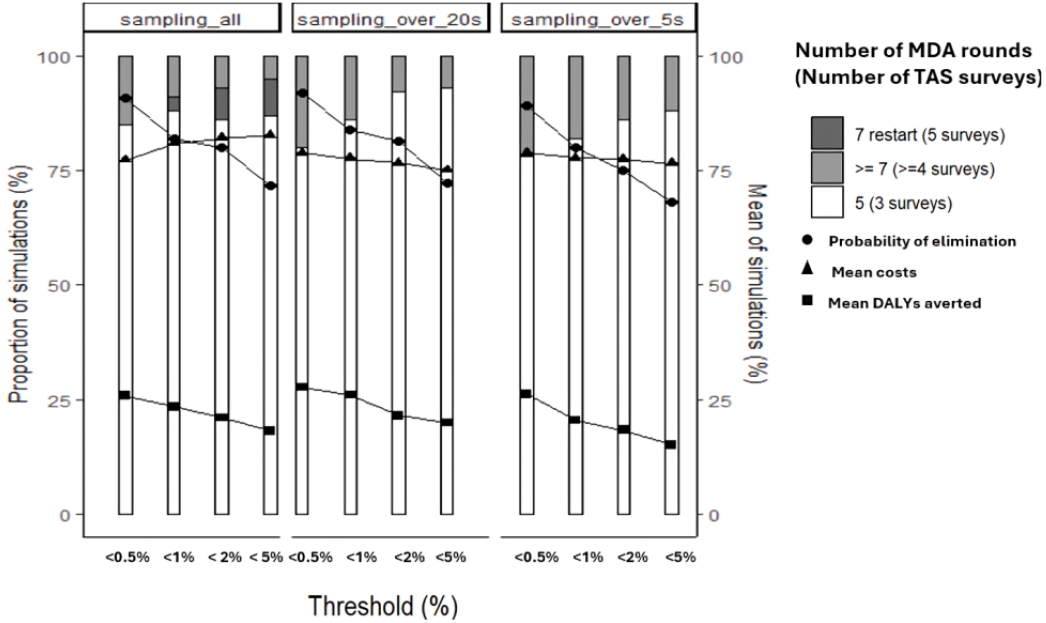
Epidemiological outcomes for different mf stopping threshold prevalences for TAS with 5-10% baseline prevalence with 80% MDA coverage of different age-groups of the eligible population post-surveillance. Here, we represent the mean outcomes by randomly sampling approximately 30 sites with 40-60 people strafied by age per site to replicate the characteristics of an evaluation unit (EU, < 500,000 people). Note that costs and DALYs averted are normalized to the same scale for improved visualization in the plots.

Additionally in Figure 3 (triangles), a lower threshold results in fewer MDA rounds and surveys due to reduced probability of restarting after stopping, hence lower costs. However, for the lowest baseline prevalence, restarting MDA is unlikely for either threshold for children and adults, with slightly higher costs for the lower threshold due to extra rounds needed. For the entire eligible population, higher threshold costs are greater due to MDA restarts. In general, more restarts occur at higher baseline prevalences and lower MDA coverage for all thresholds due to the stochastic nature of the model dynamics accounting for higher transmission and increased treatment rounds to achieve elimination (refer Tables B.4, B.5 and B.6, in Appendix B).

Figure 3 (squares) shows mean DALYs averted across different thresholds for 80% MDA coverage. Lowering the threshold results in more DALYs averted due to a reduction in worm burdens. Trends are similar for 65% MDA coverage. The choice of threshold depends on epidemiological context and economic considerations, including WTP_DALY_.

To evaluate costs, health impact, and monetization benefits of local elimination, we use expected incremental net monetary benefit (EINMB). Higher EINMB indicates optimal cost-effectiveness at a given WTP_DALY_. Our findings (Figure 4a) show that at 80% coverage, switching to a lower threshold is cost-effective across all baseline prevalences, keeping costs per DALY averted below national WTP thresholds (positive EINMB). Variability in results is due to demographic factors such as age, treatment strategy, and population growth [40]. At 65% coverage (Figure 4b), more rounds and surveys suggest switching to a lower threshold is cost-effective based on WTP per 1% increase in local elimination probability, aligning with GPELF goals (refer Tables B.7, B.8 in Appendix B). For WTPs of approximately $4200, $3000, and $1000 per 1% increase in local elimination for different baseline prevalence, switching is recommended (Figure 4b, black solid line).

**Figure 4:**
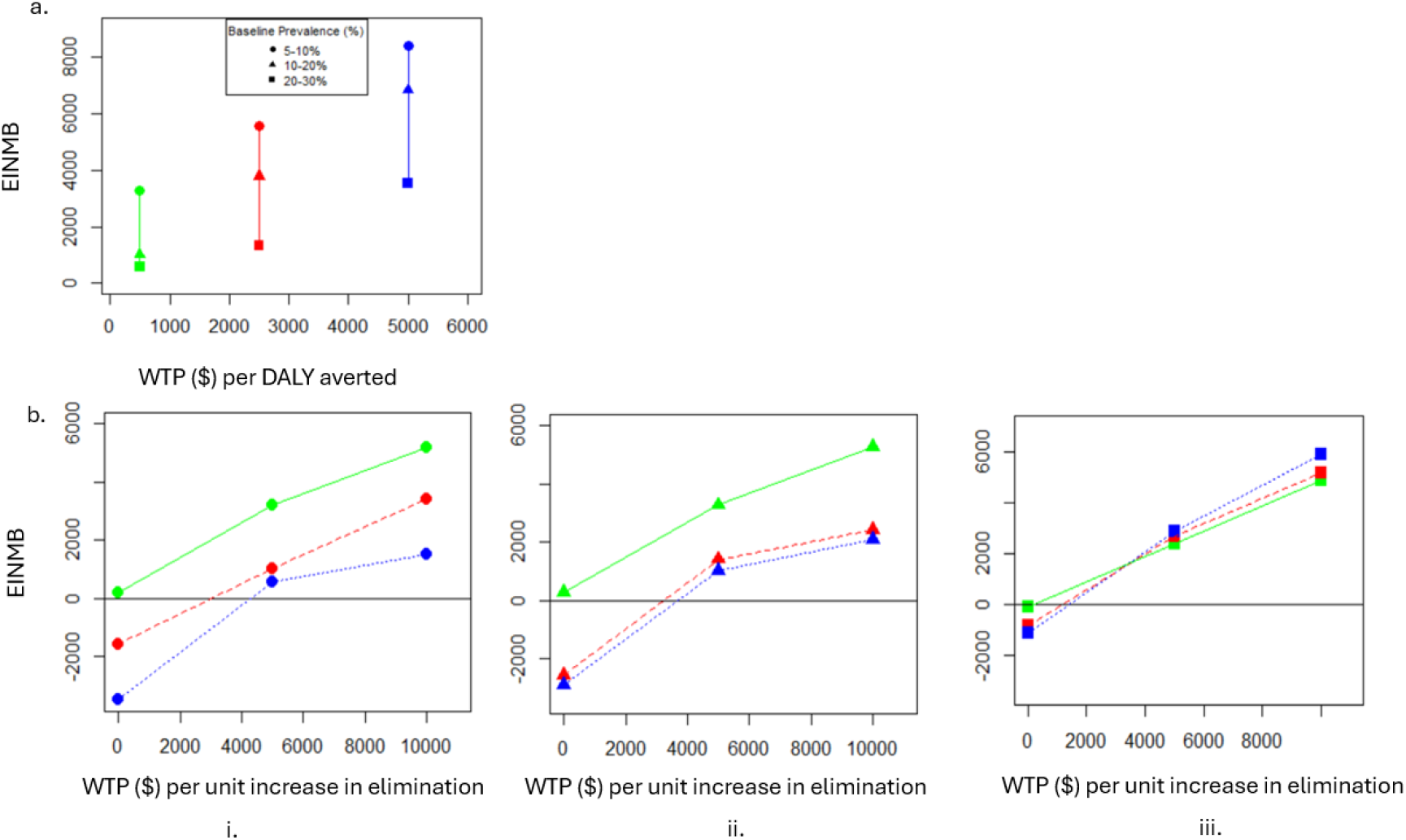
a. EINMB based on the WTP for range of DALY averted for morbidity: $500 (green), $2500 (red), $5000 (blue) for 5-10% (circles), 10-20% (triangles) and 20-30% (squares) baseline prevalence for a sample of adults. b. EINMB based on the WTP for 1% increase in probability of elimination from $0-$10,000 and the WTP_DALY_: $500 (green), $2500 (red), $5000 (blue) for sample of adults for (i) 5-10% - circles (ii) 10-20% - triangles (iii) 20-30% - squares baseline prevalences comparing <0.5% threshold in a sample of adults with respect to <1% threshold of mf prevalence in children (comparator).

Health economic decision-analytic models are used to estimate the expected net benefits of competing decision options. The true values of the input parameters of such models are rarely known with certainty, and it is often useful to quantify the value to the decision maker of reducing uncertainty through collecting new data. In the context of understanding how to measure the prevalence for different stopping threshold with precision, we need a handle to quantify uncertainty revolving around the costs due to MDA rounds and surveys alongside DALYs averted and unit increase in probability of elimination. In this light, the value of the proposed research design for every additional sample size can be quantified by the EVSI metric as defined in Section 2. In Figure 5 (a)-(b), we find that the EVSI peaks around a WTP_Elimination_ of $2,500-$3,000 (max) for <0.5% and <1% stopping threshold. On the other hand, for Figure 5 (c)-(d) we see that the EVSI peaks around a WTP_Elimination_ of about $1,500-$1,700 (max) for <2% and < 5%. This corresponds to the fact that additional information obtained from extra EVSI per person (illustrated in Figure 5 by higher peaks in 0.5% than 5%) in lower stopping thresholds is more beneficial to account for uncertainty quantification of MDA stopping decisions as justified for 5-10% baseline prevalence. Additionally, in Tables B.11, B.12 and B.13 (see Appendix B), we find that moment matching is much faster than the benchmark nested Monte-Carlo method, although both converge to the same EVSI at larger sample sizes. Figure 5 illustrates the theoretically established trend in the fact that the EVSI (dark blue lines) approached the EVPPI (thick dark blue line below EVPI) as defined in Section 2 at larger sample sizes, indicating that the value of information gained from larger studies may approach the theoretical maximum value of removing the uncertainty of reaching elimination based on the different thresholds. In order to further test the robustness of the MDA stopping decision based on the cost-effectiveness of lower stopping threshold (< 0.5% mf prevalence in adults), we rely on the Linear Wasserstein Framework in conjunction with PCA and LDA. In Figure 6a, we find the scattergram of the costs with DALYs averted with 80% MDA coverage for different baseline prevalences (represented as circles, triangles and squares) show that the lower stopping thresholds (< 0.5%) are cost-effective using the fixed country-specific WTP_DALY_ ranging from $500-$5,000 when predicted on the unseen 20% of the feature vector (test sample). Each symbol represents the mean incremental costs to incremental DALYs averted to obtain a standardized comparison to the EINMB metric in Figure 4a. Likewise, in Figure 6b. we find the scattergram of the costs with DALYs averted in addition to the probability of elimination show when <0.5% stopping thresholds are cost-effective with a narrower estimated WTP_Elimination_ per unit increase in the probability of elimination from $500-$3000 for 65% MDA coverage when predicted on the unseen 20% of the feature vector (test sample). Similarly, each symbol here represents the mean incremental costs to incremental probability for elimination to obtain a standardized comparison to the EINMB metric in Figure 4b. We remark that the results illustrated in Figure 6 a and b were computed using Step 1-3 of Algorithm 2 with 80% training sample size. Finally, in Figure 6c, we illustrate the EVSI per person using the utility gained from additional information predicted from LDA (see Algorithm 2 for more details) with the optimum WTP_Elimination_ estimated. We observe that as baseline prevalence decreases, the optimum WTP_Elimination_ to make the lower stopping threshold cost-effective increases due to more additional benefits gained from elimination and DALYs averted as demonstrated in Figure 3. Consequently, in Tables B.9 and B.10 (refer to Supplementary Appendix B), we present the classification error for accurately predicting the baseline prevalences (class labels) for different stopping thresholds which decreases as training sample sizes increase. This improvement enhances the power of the utility function, which reflects the additive benefits gained with varying stopping thresholds, as indicated by the EVSI metric. This effect is achieved by training the LDA classifier with different fractions of the sample sizes, demonstrating that larger training datasets lead to more accurate classifications and thus greater potential benefits from additional data.

**Figure 5:**
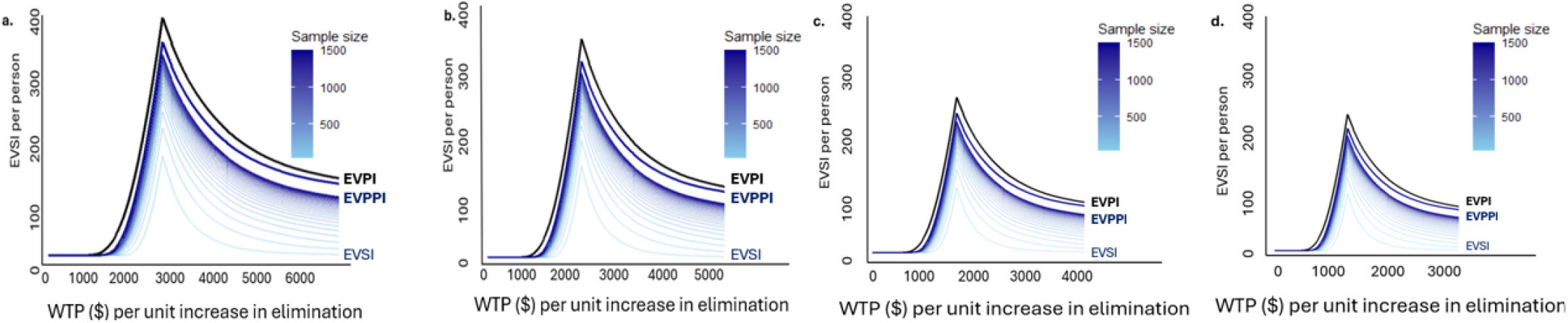
EVSI implemented using the moment matching method for different thresholds (a: <0.5%, b: <1%, c: <2% and d: <5%) to evaluate the uncertainty related to the cost-effectiveness of different stopping threshold for TAS under a range of WTP per unit increase in elimination for 5-10% baseline prevalence in a sample of adults with 65% MDA coverage.

**Figure 6:**
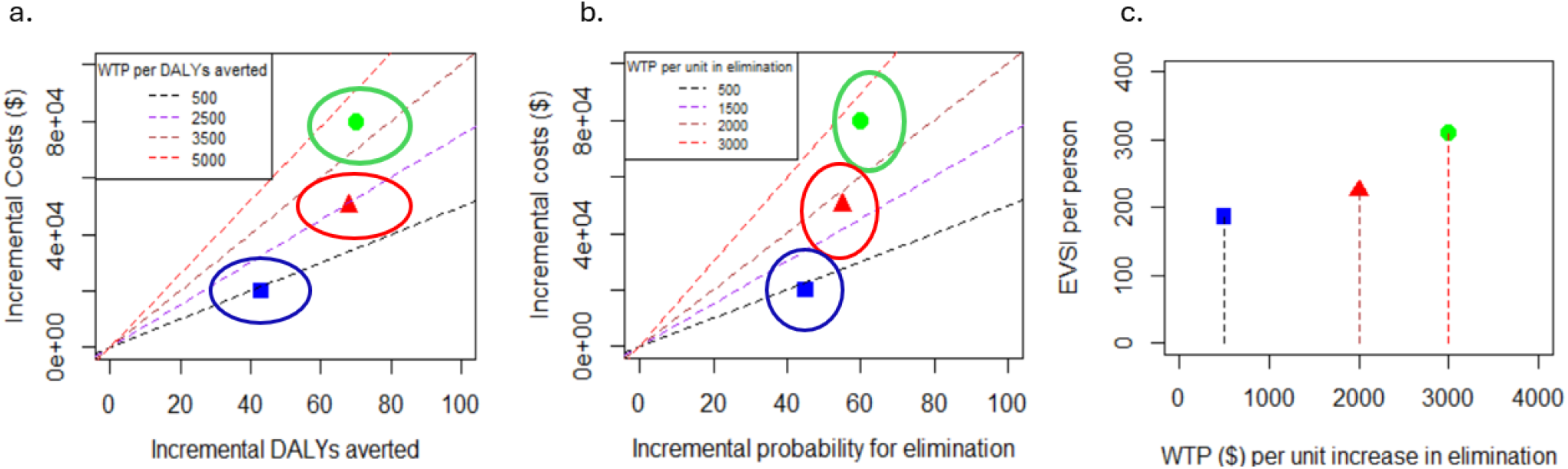
Summary figure illustrating the variations in the cost-effectiveness of the lower threshold for TAS using the Linear Wasserstein framework by considering the total costs and DALYs averted, probability for elimination as inputs. The LOT embeddings are projected into a lower-dimension space using PCA and this reduced feature vector from PCA is trained using LDA (80:20 split) for classifying <0.5% threshold at different baseline prevalence (labels) depicted as (circles) 5-10%, (triangles) 10-20% and (squares) 20-30% alongside the WTP for DALY averted for morbidity: $500 (green), $2,500 (red), $5,000 (blue) for a sample of adults. a. Scattergram of incremental costs and DALYs averted for 80% MDA coverage b. Scattergram of incremental costs and unit increase in probability of elimination for 65% MDA coverage c. EVSI per person for the estimated optimum WTP per unit increase in elimination. Note: The comparator chosen as the reference template is <1% mf prevalence as the stopping threshold for TAS in children. The ellipses estimated from the covariance matrix and the mean vectors of each baseline prevalence (class labels) denote the 95% confidence intervals accounting for the uncertainty and variability within the distribution of each class. Each symbol represents the mean of the incremental costs to DALYs averted or probability of elimination classified by their respective baseline prevalences.

## 6. Discussion

The probability of local elimination is determined by stopping thresholds, which are crucial for many disease control policies. That being said, it would be worthwhile to look into the effects of a lower threshold on program costs overall as well as whether it raises the likelihood of local elimination. The application of such a lower threshold in China [57] and its significance in effective LF control serve as examples of the potential advantages of a lower threshold, which this study highlights. However, the GPELF can use this example to gather crucial data in order to establish standards for assessing whether MDA has been successful in bringing the infection prevalence down to a point where recrudescence is unlikely to happen.

As we reduce the mf prevalence threshold from <1% to <0.5%, the likelihood of local elimination increases, according to our analysis of the effects of various stopping thresholds for TAS across 30 sites. Diminished DALY burden and restart probability are mitigated by a lower threshold, despite requiring more rounds. Employing the defined EINMB metric for CEA reveals that switching to a lower threshold is economical at 80% MDA coverage. However, for 65% MDA coverage, extra benefits are needed, such as utilizing the WTP_Elimination_ per unit increase for elimination. The low amount of data, especially on systematic non-adherence and wider disease impacts like mental illness, is the reason for the conservative morbidity estimates [49; 30].

The expanded use of CEA in healthcare faces several challenges. First, decision-makers must account for social concerns like prioritizing the sick, reducing health disparities by integrating more social concerns into CEA techniques. Second, current CEA practices, focused on evaluating new strategies or technologies, often overlook signs of resource misallocation. Third, assessing the broad range of interventions needed for CEA to improve allocative efficiency can be prohibitively expensive and time-consuming. Additionally, many CEA studies produce context-specific results, limiting their applicability to different populations. Progress towards providing timely, affordable information on the costs and effects of various interventions remains limited, particularly for LMIC [48; 60].

On the other hand, the Linear Wasserstein Framework despite being mathematically rigorous has its own limitations. Firstly, the framework makes several modelling assumptions. Namely, that the distance should be proportional to the cost of translations. This can make the distance sensitive to outliers. Secondly, the Linear Wasserstein distance is also an approximation of the Wasserstein distance and this approximation may deteriorate depending on the local curvature. Thirdly, being balanced, this framework requires equal number of datapoints (1000) for each probability measures derived from each of the pointcloud data for the different baseline prevalences (classes) which can disproportionately affect the robustness of the results, if class-imbalance exists. A further extension to this framework could be to generate future projections of the model simulations for different baseline prevalences using fewer runs to save the computational power of the TRANSFIL model from a Bayesian perspective [37] or use state-of-art methods such as graph-based semi-supervised methods [9] that can leverage the advantages of this geometric embedding when very little information on the data is provided so that it can learn the geometry of the underlying point cloud data effectively.

Our study assumes constant survey implementation costs, excluding potential out-of-pocket expenses and future cost changes [41]. Despite challenges in estimating precise costs for MDA and TAS due to incomplete records and data access issues, simulations help understand the TAS threshold’s impact on stopping MDA. A limitation in this study is the exclusion of vector control benefits, which remain debated. While some studies suggest combined MDA and vector control benefits in low endemic regions, others find no added advantage over MDA alone upon which further research is needed. Another major limitation is that our modelling study relies on *Culex* vector due to its increased efficiency in transmission. Although direct implication of *Culex* species in the transmission of LF in West and Central Africa is still not well documented [42; 4], in East Africa, *Culex* species particularly *Cx. quinquefasciatus* is known to have a major role in LF transmission [17; 33]. With changing climate associated to increased traffic between East and West African countries and rapid expansion of this species in urban settings, it is becoming crucial to assess the role of *Culex* species in the transmission of diseases like LF. We also restrict our analysis to the IA drug, but studies [50] for oncho have found that IA may not lead to elimination of transmission (EoT) in all endemic areas and moxidectin-based strategies could accelerate progress toward EoT and reduce programmatic delivery costs compared with ivermectin-based strategies. We also note that the use of the three-drug combination IDA, has particular challenges [45] for survey design due to reductions in mf density but not Ag over one or two rounds of treatment [27].

Despite these drawbacks, our research emphasizes how important it is to choose the right framework for uncertainty quantification when making decisions, especially when it comes to disease interventions, particularly LF. It is also essential to comprehend the dynamics of local elimination post-threshold crossing and how it interacts with LF interventions. Our research indicates that although there is a long transient phase involved in the path to LF local elimination post-MDA surveillance, lower thresholds may help programs achieve their objectives. In addition, we also propose the need for better framework to quantify the uncertainty inherent in the model parameters to analyze the cost-effectiveness of lowering the stopping threshold in LF.

## Data Availability

All data produced in the present study are available upon reasonable request to the authors

## 9. CRediT authorship contribution statement

Mary Chriselda Antony Oliver: Conceptualization, Methodology, Software, Formal analysis, Writing-original draft, Writing-review and editing, Visualization. Matthew Graham: Software, Writing-review and editing. Ioanna Manolopoulou, Graham F. Medley, Lorenzo Pellis - Writing-review and editing. Koen B Poewels, Matthew Thorpe, T. Deirdre Hollingsworth - Conceptualization, Methodology, Writing - review and editing, Supervision.

