## Supplementary Material for "Uncertainty Quantification in Cost-effectiveness Analysis for Stochastic-based Infectious Disease Models: Insights from Surveillance on Lymphatic Filariasis"

### Appendix A. Algorithms

**Algorithm 1** Moment Matching Algorithm for Estimating EVSI

**Input:** Number of outer samples $K$, number of inner samples $L$
**Output:** Estimated EVSI
Initialize $\mathrm{NMB}_{\mathrm{outer}} \leftarrow 0$ and $\mathrm{NMB}_{\mathrm{baseline}} \leftarrow 0$
**for** $k = 1$ to $K$ **do**
  Sample $\theta_k$ from the prior distribution $p(\theta)$
  Generate data $x_k$ from the likelihood $p(X|\theta = \theta_k)$
  Approximate the posterior $p(\theta|x_k)$ using moment matching:
  Compute the sample mean $\hat{\mu}_k$ and sample covariance $\hat{\Sigma}_k$ of the posterior samples $\theta_{l,k}$ given $x_k$
  $\mathrm{NMB}_{\mathrm{inner},k} \leftarrow 0$
  **for** $l = 1$ to $L$ **do**
    Sample $\theta_{l,k}$ from $\mathcal{N}(\hat{\mu}_k, \hat{\Sigma}_k)$
    Compute $\mathrm{NMB}(j, \theta_{l,k}|\mathrm{WTP}_{\mathrm{DALY}}, \mathrm{WTP}_{\mathrm{Elimination}})$ for each $j = \{1, 2, \cdots, J\}$
    $\mathrm{NMB}_{\mathrm{inner},k} \leftarrow \mathrm{NMB}_{\mathrm{inner},k} + \frac{1}{L} \max_{j=\{1,2,\cdots,J\}} \mathrm{NMB}(j, \theta_{l,k})$
  **end for**
  $\mathrm{NMB}_{\mathrm{outer}} \leftarrow \mathrm{NMB}_{\mathrm{outer}} + \frac{1}{K}\mathrm{NMB}_{\mathrm{inner},k}$
**end for**
**Estimate the maximum expected NMB without additional data:**
**for** $i = 1$ to $K$ **do**
  Sample $\theta_i$ from the prior distribution $p(\theta)$
  Compute $\mathrm{NMB}(j, \theta_i|\mathrm{WTP}_{\mathrm{DALY}}, \mathrm{WTP}_{\mathrm{Elimination}})$ for each $j = \{1, 2, \cdots, J\}$
  $\mathrm{NMB}_{\mathrm{baseline}} \leftarrow \mathrm{NMB}_{\mathrm{baseline}} + \frac{1}{K} \max_{j=\{1,2,\cdots,J\}} \mathrm{NMB}(j, \theta_i)$
**end for**
**Compute EVSI:**
$\hat{\mathrm{EVSI}} = \mathrm{NMB}_{\mathrm{outer}} - \mathrm{NMB}_{\mathrm{baseline}}$
**Return** $\hat{\mathrm{EVSI}}$

**Algorithm 2** Compute EVSI using LOT + PCA + LDA

**Input:** Data matrices $\{\mu_i\}_{i=1}^N$, Reference measure $\sigma$, Class labels $\{y_i\}_{i=1}^N$, Number of PCA components $p$, Number of simulated datasets $M$, Utility function $U(\hat{y}_j, y_j)$ such that $U(\hat{y}_j, y_j) = 1$ if $\hat{y}_j = y_j$, else 0.
**Output:** EVSI estimate
**Step 1: Compute the Linear Optimal Transport (LOT)**
**for** $i = 1$ to $N$ **do**
  Compute optimal transport map $T_i^*$ between $\sigma$ and $\mu_i$ such that:

$$d_{W^p}(\mu_i, \sigma) := \left( \int_\Omega |x - T_i^*(x)|^p \, \mathrm{d}\sigma(x) \right)^{\frac{1}{p}}$$

  Compute the projections $P(\mu_i) = T_i^* - \mathrm{Id}$, where Id is the identity map.
**end for**
**Step 2: Apply PCA on the projections**
Construct matrix $\mathbf{P} \in \mathbb{R}^{N \times d}$ where each row is $\{P(\mu_i)\}_{i=1}^N$.
Center the data: $\mathbf{P}_c \leftarrow \mathbf{P} - \mathbb{E}(\mathbf{P})$.
Compute the covariance matrix: $\mathbf{C} \leftarrow \frac{1}{N}\mathbf{P}_c^T\mathbf{P}_c$
Compute eigenvalues and eigenvectors of $\mathbf{C}$
Select top $p$ dominant eigenvectors to form the PCA projection matrix $\mathbf{P}_{\mathrm{PCA}} \in \mathbb{R}^{d \times p}$
Project the centered data onto PCA components: $\mathbf{X} = \mathbf{P}_c\mathbf{P}_{\mathrm{PCA}}$.
**Step 3: Train the LDA classifier**
Train the LDA classifier once using the PCA components for different $L = 20\%, 40\%, 60\%, 80\%$ of the data i.e., $\{\mathbf{X}_i\}_{i=1}^L$ with the class labels $\{y_i\}_{i=1}^L$ where $L$ is the size of the training dataset.
**Return** Predicted labels $\{\hat{y}_j\}_{j=1}^V$ on the test data $\{\mathbf{X}_j\}_{j=1}^V$ where $V = N - L$ is the size of the test dataset. Decision boundary giving the estimate for $\mathrm{WTP}_{\mathrm{Elimination}}$ for 65% MDA coverage.
**Step 4: Evaluate Utility function**
Compute expected utility $\mathrm{EU}_{\mathrm{current}} = \frac{1}{V}\sum_{j=1}^V U(\hat{y}_j, y_j)$.
**Step 5: Train on the Augmented Dataset**
**for** $k = 1$ to $M$ **do**
  Generate bootstrap samples from the LOT projections $\{P(\mu_i)\}_{i=1}^N$, namely $\{P'(\mu_i)\}_{i=1}^N$.
  Follow Steps 2-3 and compute the PCA components for the new bootstrap sample resulting in $\{\mathbf{X}'_i\}_{i=1}^L$ with class labels $\{y'_i\}_{i=1}^L$. Train LDA on this bootstrapped dataset for $L = 20\%, 40\%, 60\%, 80\%$.
  Obtain predicted class labels $\{\hat{y}'_j\}_{j=1}^V$ for test data $\{\mathbf{X}'_j\}_{j=1}^V$ where $V = N - L$.
  Compute expected utility for the $k$-th bootstrap sample: $\mathrm{EU}_{\mathrm{new},k} = \frac{1}{V}\sum_{j=1}^V U(\hat{y}'_j, y'_j)$.
**end for**
**Step 6: Compute EVSI**
Compute average expected utility with additional data: $\mathrm{EU}_{\mathrm{sample}} = \frac{1}{M}\sum_{k=1}^M \mathrm{EU}_{\mathrm{new},k}$.
Compute EVSI: $\mathrm{EVSI} = \mathrm{EU}_{\mathrm{sample}} - \mathrm{EU}_{\mathrm{current}}$.
**Return** EVSI

### Appendix B. Supplementary Material: Additional Results

Table B.1: Probability of Elimination for different stopping thresholds for TAS sampling for people aged $\geq$ 20 years (adults)

| | | Probability of Elimination for different stopping thresholds for TAS | | | |
|---|---|---|---|---|---|
| MDA coverage | Mean baseline prevalence | (<0.5%) | (<1%) | (<2%) | (<5%) |
| | 5-10% | 91.8% (90.2 - 92.28) | 83.8% (80.97 - 84.8) | 81.47% (80.23 - 82.27) | 72.16% (70.67 - 72.28) |
| 80% | 10-20% | 77.28% (75.56 - 78.23) | 71.03% (70.03 - 68.97) | 65.88% (64.26 - 67.28) | 61.78% (60.23 - 62.97) |
| | 20-30% | 67.68% (63.24 - 68.52) | 65.91% (64.06 - 67.28) | 63.96% (61.34 - 65.34) | 61.51% (60.23 - 63.23) |
| | 5-10% | 90.7% (88.13 - 92.02) | 87.65% (85.34 - 89.70) | 85.43% (82.02 - 89.22) | 80.03% (79.89 - 82.27) |
| 65% | 10-20% | 75.12% (72.02 - 77.36) | 70.10% (69.89 - 72.81) | 65.76% (64.83 - 67.86) | 60.47% (58.96 - 62.21) |
| | 20-30% | 65.63% (62.54 - 68.35) | 62.73% (60.83 - 69.43) | 60.15% (59.24 - 62.35) | 59.24% (54.10 - 62.37) |

Table B.2: Probability of Elimination for different stopping thresholds for TAS sampling for people aged $\geq$ 5 years (including, children)

| | | Probability of Elimination for different stopping thresholds for TAS | | | |
|---|---|---|---|---|---|
| MDA coverage | Mean baseline prevalence | (<0.5%) | (<1%) | (<2%) | (<5%) |
| | 5-10% | 89.2% (87.22 - 91.23) | 80.05% (79.23 - 81.28) | 75.04% (72.28 - 78.97) | 68.08% (65.07 - 67.83) |
| 80% | 10-20% | 72.63% (70.86 - 77.53) | 62.04% (60.23 - 63.27) | 57.33% (54.62 - 59.90) | 52.49% (50.23 - 56.24) |
| | 20-30% | 55.99% (51.02 - 56.87) | 53.5% (50.23 - 58.75) | 50.5% (48.23 - 52.55) | 48.23% (45.23 - 50.12) |
| | 5-10% | 85.23% (82.78 - 89.23) | 78.23% (76.89-80.23) | 72.25% (70.02 - 75.23) | 65.13% (62.34-68.23) |
| 65% | 10-20% | 72.15% (71.03 - 75.45) | 60.23% (58.89 - 63.23) | 55.13% (52.82-57.86) | 51.23% (50.96 - 56.87) |
| | 20-30% | 55.23% (50.54 - 58.35) | 51.23% (50.83 - 56.43) | 48.23% (46.24 - 50.35) | 46.73% (44.78 - 48.87) |

Table B.3: Probability of Elimination for different stopping thresholds for TAS sampling for everyone eligible

| | | Probability of Elimination for different stopping thresholds for TAS | | | |
|---|---|---|---|---|---|
| MDA coverage | Mean baseline prevalence | (<0.5%) | (<1%) | (<2%) | (<5%) |
| | 5-10% | 90.7% (87.23 - 92.18) | 81.76% (80.17 - 83.63) | 80.07% (78.86 - 82.27) | 71.58% (70.57 - 72.28) |
| 80% | 10-20% | 74.57% (75.56 - 78.23) | 71.03% (70.03 - 68.97) | 65.88% (64.26 - 67.28) | 61.78% (60.23 - 62.97) |
| | 20-30% | 60.08% (59.84 - 62.52) | 57.64% (55.23 - 60.12) | 54.23% (51.24 - 57.98) | 50.23% (49.93 - 53.29) |
| | 5-10% | 88.13% (83.23 - 90.02) | 78.23% (75.34-79.70) | 70.24% (69.75 - 73.25) | 68.23% (65.89-70.87) |
| 65% | 10-20% | 72.13% (70.02-77.36) | 65.52% (60.23 - 68.87) | 60.23% (59.83-62.86) | 58.23% (55.86 - 60.81) |
| | 20-30% | 58.23% (57.04 - 60.05) | 55.23% (54.03 - 57.89) | 52.12% (50.04 - 55.85) | 48.23% (46.23 - 50.91) |

Table B.4: Total MDA rounds with TAS surveys for different MDA coverage levels sampling for people aged ≥ 20 years and above.

| | | Total MDA rounds (TAS surveys) | | | | | | | |
|---|---|---|---|---|---|---|---|---|---|
| Mean baseline prevalence | Total Number of MDA rounds (TAS surveys) | (<0.5%) | | (<1%) | | (<2%) | | (<5%) | |
| | MDA coverage | 80% | 65% | 80% | 65% | 80% | 65% | 80% | 65% |
| 5-10% | 5 rounds (3 surveys) | 80 | 76 | 86 | 80 | 92 | 88 | 93 | 90 |
| | ≥ 7 rounds (4 surveys) | 20 | 24 | 14 | 20 | 8 | 12 | 7 | 10 |
| | 7 restart (5 surveys) | 0 | 0 | 0 | 0 | 0 | 0 | 0 | 0 |
| 10-20% | 5 rounds (3 surveys) | 82 | 70 | 88 | 72 | 90 | 77 | 88 | 82 |
| | ≥ 7 rounds (4 surveys) | 18 | 26 | 12 | 24 | 10 | 17 | 9 | 12 |
| | 7 restart (5 surveys) | 0 | 4 | 0 | 4 | 0 | 6 | 3 | 6 |
| 20-30% | 5 rounds (3 surveys) | 45 | 35 | 43 | 38 | 44 | 39 | 46 | 41 |
| | ≥ 7 rounds (4 surveys) | 50 | 57 | 50 | 55 | 46 | 54 | 45 | 48 |
| | 7 restart (5 surveys) | 5 | 8 | 7 | 7 | 10 | 7 | 9 | 11 |

Table B.5: Total MDA rounds with TAS surveys for different MDA coverage levels sampling for people aged ≥ 5 years and above.

| | | Total MDA rounds (TAS surveys) | | | | | | | |
|---|---|---|---|---|---|---|---|---|---|
| Mean baseline prevalence | Total Number of MDA rounds (TAS surveys) | (<0.5%) | | (<1%) | | (<2%) | | (<5%) | |
| | MDA coverage | 80% | 65% | 80% | 65% | 80% | 65% | 80% | 65% |
| 5-10% | 5 rounds (3 surveys) | 78 | 74 | 82 | 80 | 86 | 82 | 88 | 84 |
| | ≥ 7 rounds (4 surveys) | 22 | 26 | 18 | 20 | 14 | 20 | 12 | 0 |
| | 7 restart (5 surveys) | 0 | 0 | 0 | 0 | 0 | 0 | 0 | 0 |
| 10-20% | 5 rounds (3 surveys) | 82 | 68 | 85 | 70 | 87 | 77 | 88 | 80 |
| | ≥ 7 rounds (4 surveys) | 18 | 28 | 15 | 26 | 13 | 18 | 8 | 13 |
| | 7 restart (5 surveys) | 0 | 4 | 0 | 4 | 0 | 5 | 4 | 7 |
| 20-30% | 5 rounds (3 surveys) | 37 | 36 | 42 | 39 | 41 | 41 | 40 | 43 |
| | ≥ 7 rounds (4 surveys) | 54 | 52 | 46 | 52 | 43 | 46 | 42 | 42 |
| | 7 restart (5 surveys) | 9 | 12 | 12 | 9 | 16 | 13 | 18 | 15 |

Table B.6: Total MDA rounds with TAS surveys for different MDA coverage levels sampling everone eligible in the population

| | | Total MDA rounds (TAS surveys) | | | | | | | |
|---|---|---|---|---|---|---|---|---|---|
| Mean baseline prevalence | Total Number of MDA rounds (TAS surveys) | (<0.5%) | | (<1%) | | (<2%) | | (<5%) | |
| | MDA coverage | 80% | 65% | 80% | 65% | 80% | 65% | 80% | 65% |
| 5-10% | 5 rounds (3 surveys) | 85 | 80 | 88 | 82 | 86 | 83 | 87 | 85 |
| | ≥ 7 rounds (4 surveys) | 15 | 20 | 9 | 12 | 7 | 9 | 5 | 9 |
| | 7 restart (5 surveys) | 0 | 0 | 3 | 6 | 7 | 8 | 8 | 6 |
| 10-20% | 5 rounds (3 surveys) | 78 | 63 | 76 | 65 | 77 | 68 | 80 | 72 |
| | ≥ 7 rounds (4 surveys) | 18 | 32 | 18 | 28 | 15 | 23 | 11 | 17 |
| | 7 restart (5 surveys) | 4 | 5 | 6 | 7 | 8 | 9 | 9 | 11 |
| 20-30% | 5 rounds (3 surveys) | 41 | 35 | 40 | 36 | 42 | 42 | 44 | 41 |
| | ≥ 7 rounds (4 surveys) | 52 | 59 | 51 | 55 | 47 | 49 | 43 | 47 |
| | 7 restart (5 surveys) | 7 | 6 | 9 | 9 | 11 | 9 | 13 | 12 |

Table B.7: : Expected incremental net monetary benefit (in dollars) of switching from <1% threshold to <0.5% threshold in the TAS for each setting for a sample of adults aged 20 years and above. Note: The comparator for computing the EINMB is the <1% threshold in people aged 5 years and above for the individual baseline prevalence and MDA coverage of the total population. This has been chosen for a range of WTP for DALY averted and probability of elimination so that we can estimate the cost-effectiveness of the threshold at different known baseline prevalence and MDA coverage.

| | (WTP per DALY averted) | | | | | |
|---|---|---|---|---|---|---|
| Baseline Prevalence | $ 500 | | $2500 | | $5000 | |
| | MDA Coverage | | MDA Coverage | | MDA Coverage | |
| | 80% | 65% | 80% | 65% | 80% | 65% |
| Willingness to pay $0 per 1% increase in the probability of elimination to switch thresholds | | | | | | |
| 5-10% | $3,286 | $189 | $1,009 | -$1589 | $5,79 | -$3503 |
| 10-20% | $5,575 | $275.32 | $3,781 | -$2589 | $1,334 | -$2892 |
| 20-30% | $8,373 | -$102 | $6,823 | -$803 | $3,534 | -$1106 |
| Willingness to pay $10,000 per 1% increase in the probability of elimination to switch thresholds | | | | | | |
| 5-10% | $3,286 | $5189 | $1,009 | $3,411 | $5,79 | $1497 |
| 10-20% | $5,575 | $5275 | $3,781 | $2,411 | $1,334 | $2108 |
| 20-30% | $8,373 | $4898 | $6,823 | $5198 | $3,534 | $5898 |
| Minimum willingness to pay $5,000 per 1% increase in probability of elimination to switch thresholds | | | | | | |
| 5-10% | $3,286 | $3189 | $1,009 | $1,411 | $5,79 | $497 |
| 10-20% | $5,575 | $3275 | $3,781 | $1411 | $1,334 | $1,008 |
| 20-30% | $8,373 | $2398 | $6,823 | $2197 | $3,534 | $2,894 |

Table B.8: Expected incremental net monetary benefit (in dollars) of switching from <1% threshold to <0.5% threshold in the TAS for each setting when everyone eligible in the population has been considered. Note: The comparator for computing the EINMB is the <1% threshold in people aged 5 years and above for the individual baseline prevalence and MDA coverage of the total population. This has been chosen for a range of WTP for DALY averted and probability of elimination so that we can estimate the cost-effectiveness of the threshold at different known baseline prevalence and MDA coverage.

| | (WTP per DALY averted) | | | | | |
|---|---|---|---|---|---|---|
| Baseline Prevalence | $ 500 | | $2500 | | $5000 | |
| | MDA Coverage | | MDA Coverage | | MDA Coverage | |
| | 80% | 65% | 80% | 65% | 80% | 65% |
| Willingness to pay $0 per 1% increase in the probability of elimination to switch thresholds | | | | | | |
| 5-10% | $4,398 | $979 | $2,469 | -$2569 | $8,29 | -$3593 |
| 10-20% | $6,695 | $459 | $5281 | -$2793 | $1,524 | -$3092 |
| 20-30% | $9,203 | -$112 | $7,243 | -$1243 | $4,864 | -$836 |
| Willingness to pay $10,000 per 1% increase in the probability of elimination to switch thresholds | | | | | | |
| 5-10% | $4,398 | $5979 | $2,469 | $2,431 | $8,29 | $1407 |
| 10-20% | $6,695 | $5459 | $5281 | $2,207 | $1,524 | $1907 |
| 20-30% | $9,203 | $4888 | $7,243 | $3757 | $4,864 | $4164 |
| Minimum willingness to pay $5,000 per 1% increase in probability of elimination to switch thresholds | | | | | | |
| 5-10% | $4,398 | $3979 | $2,469 | $431 | $8,29 | $407 |
| 10-20% | $6,695 | $3459 | $5281 | $207 | $1,524 | $9,08 |
| 20-30% | $9,203 | $2888 | $7,243 | $1757 | $4,864 | $3,164 |

Table B.9: Classification error on test data using LDA for different values of the training sample size and different thresholds using point cloud data of costs and DALYs averted using fixed WTP for DALYs averted for 80% MDA coverage for sample of adults.

| Baseline Prevalence | Number of Training Sample | Classification Error on test data | | | |
|---|---|---|---|---|---|
| | | 0.5% | 1% | 2% | 5% |
| 5-10% | 20% | 0.2 | 0.27 | 0.35 | 0.37 |
| | 50% | 0.17 | 0.21 | 0.27 | 0.32 |
| | 60% | 0.10 | 0.12 | 0.19 | 0.23 |
| | 80% | 0.05 | 0.07 | 0.09 | 0.12 |
| 10-20% | 20% | 0.42 | 0.46 | 0.49 | 0.51 |
| | 50% | 0.37 | 0.43 | 0.45 | 0.48 |
| | 60% | 0.32 | 0.35 | 0.37 | 0.39 |
| | 80% | 0.21 | 0.24 | 0.29 | 0.31 |
| 20-30% | 20% | 0.59 | 0.64 | 0.67 | 0.72 |
| | 50% | 0.52 | 0.43 | 0.37 | 0.35 |
| | 60% | 0.32 | 0.35 | 0.39 | 0.42 |
| | 80% | 0.33 | 0.37 | 0.42 | 0.44 |

Table B.10: Classification error on test data using LDA for different values of the training sample size and different thresholds using a point cloud of costs with DALYs averted, Probability of elimination using estimated WTP for 1% increase in probability of elimination for 65% MDA coverage using a sample of adults.

| Baseline Prevalence | Number of Training Sample | Classification Error on the test data | | | |
|---|---|---|---|---|---|
| | | 0.5% | 1% | 2% | 5% |
| 5-10% | 20% | 0.3 | 0.34 | 0.37 | 0.41 |
| | 50% | 0.27 | 0.31 | 0.35 | 0.39 |
| | 60% | 0.23 | 0.27 | 0.29 | 0.31 |
| | 80% | 0.15 | 0.17 | 0.19 | 0.22 |
| 10-20% | 20% | 0.43 | 0.47 | 0.52 | 0.57 |
| | 50% | 0.41 | 0.43 | 0.47 | 0.52 |
| | 60% | 0.37 | 0.39 | 0.41 | 0.43 |
| | 80% | 0.35 | 0.37 | 0.39 | 0.41 |
| 20-30% | 20% | 0.56 | 0.61 | 0.63 | 0.62 |
| | 50% | 0.42 | 0.43 | 0.53 | 0.57 |
| | 60% | 0.35 | 0.37 | 0.41 | 0.47 |
| | 80% | 0.27 | 0.33 | 0.37 | 0.44 |

Table B.11: Comparison of the EVSI per person for Moment Matching (MM), Nested Monte Carlo (MC) and LOT+PCA+LDA methods for 65% MDA coverage in 5-10% mf baseline prevalence

| Threshold (%) | Sample Size (500) | | | Sample Size (1000) | | | Sample Size (1500) | | |
|---|---|---|---|---|---|---|---|---|---|
| | MC | MM | LOT+LDA | MC | MM | LOT+LDA | MC | MM | LOT+LDA |
| EVSI per Person | | | | | | | | | |
| <0.5% | 250 | 200 | 220 | 285 | 270 | 276 | 295 | 300 | 310 |
| <1% | 210 | 180 | 185 | 263 | 250 | 255 | 277 | 280 | 287 |
| <2% | 152 | 130 | 145 | 185 | 170 | 165 | 217 | 220 | 229 |
| <5% | 120 | 100 | 110 | 155 | 140 | 145 | 175 | 180 | 192 |
| Computational Time | | | | | | | | | |
| <0.5% | >8 hrs | ≈2 min | ≈1 min | >8 hrs | ≈2 min | ≈1 min | >8 hrs | ≈2 min | ≈1 min |
| <1% | >8 hrs | ≈2 min | ≈1 min | >8 hrs | ≈2 min | ≈1 min | >8 hrs | ≈2.5 min | ≈1 min |
| <2% | >8 hrs | ≈2 min | ≈1 min | >8 hrs | ≈2 min | ≈2 min | >8 hrs | ≈2 min | ≈2 min |
| <5% | >8 hrs | ≈2.5 min | ≈2 min | >8 hrs | ≈3 min | ≈2 min | >8 hrs | ≈3 min | ≈2 min |

Table B.12: Comparison of the EVSI per person for Moment Matching (MM), Nested Monte Carlo (MC) and LOT+PCA+LDA methods for 65% MDA coverage in 10-20% mf baseline prevalence

| Threshold (%) | Sample Size (500) | | | Sample Size (1000) | | | Sample Size (1500) | | |
|---|---|---|---|---|---|---|---|---|---|
| | MC | MM | LOT+LDA | MC | MM | LOT+LDA | MC | MM | LOT+LDA |
| EVSI per Person | | | | | | | | | |
| <0.5% | 197 | 175 | 185 | 245 | 230 | 237 | 272 | 270 | 275 |
| <1% | 182 | 163 | 173 | 222 | 210 | 215 | 225 | 220 | 227 |
| <2% | 140 | 120 | 134 | 162 | 160 | 175 | 175 | 170 | 172 |
| <5% | 105 | 80 | 98 | 125 | 110 | 119 | 157 | 170 | 153 |
| Computational Time | | | | | | | | | |
| <0.5% | >8 hrs | ≈2 min | ≈1 min | >8 hrs | ≈2 min | ≈1 min | >8 hrs | ≈2 min | ≈1 min |
| <1% | >8 hrs | ≈2 min | ≈1 min | >8 hrs | ≈2 min | ≈1 min | >8 hrs | ≈2.5 min | ≈1 min |
| <2% | >8 hrs | ≈2 min | ≈1 min | >8 hrs | ≈2 min | ≈2 min | >8 hrs | ≈2 min | ≈2 min |
| <5% | >8 hrs | ≈2.5 min | ≈2 min | >8 hrs | ≈3 min | ≈2 min | >8 hrs | ≈3 min | ≈2 min |

Table B.13: Comparison of the EVSI per person for Moment Matching (MM), Nested Monte Carlo (MC) and LOT+PCA+LDA methods for 65% MDA coverage in 20-30% mf baseline prevalence

| Threshold (%) | Sample Size (500) | | | Sample Size (1000) | | | Sample Size (1500) | | |
|---|---|---|---|---|---|---|---|---|---|
| | MC | MM | LOT+LDA | MC | MM | LOT+LDA | MC | MM | LOT+LDA |
| EVSI per Person | | | | | | | | | |
| <0.5% | 165 | 157 | 160 | 182 | 175 | 177 | 192 | 180 | 185 |
| <1% | 115 | 103 | 109 | 123 | 110 | 115 | 130 | 125 | 127 |
| <2% | 95 | 87 | 92 | 105 | 93 | 100 | 120 | 110 | 117 |
| <5% | 63 | 52 | 57 | 87 | 75 | 82 | 107 | 98 | 105 |
| Computational Time | | | | | | | | | |
| <0.5% | >8 hrs | ≈2 min | ≈1 min | >8 hrs | ≈2 min | ≈1 min | >8 hrs | ≈2 min | ≈1 min |
| <1% | >8 hrs | ≈2 min | ≈1 min | >8 hrs | ≈2 min | ≈1 min | >8 hrs | ≈2.5 min | ≈1 min |
| <2% | >8 hrs | ≈2 min | ≈1 min | >8 hrs | ≈2 min | ≈2 min | >8 hrs | ≈2 min | ≈2 min |
| <5% | >8 hrs | ≈2.5 min | ≈2 min | >8 hrs | ≈3 min | ≈2 min | >8 hrs | ≈3 min | ≈2 min |

### Appendix C. Supplementary Material : Transmission Model Details

The codes used in this paper, including the R version of the TRANSFIL model is available from `https://github.com/mca52/Codes_JTB.git`.

Table C.14: Table with the parameters used for TRANSFIL model.

| Parameter | Value/Scenario |
|---|---|
| Baseline mf prevalence (%) | 5 -10%, 10-20%, 20-30% |
| Drug | IA |
| MDA frequency | Annual |
| MDA coverage | 65%, 80% |
| MDA systematic non-adherence correlation | 0.2806 - 0.5351 |
| EPHP threshold (mf prevalence) | <0.5%,<1%,<2%,<5% |
| Primary vector species | Culex |
| Bite risk aggregation parameter, ($k$) | 0.01 – 0.1 |
| Annual biting rate (ABR) | 0 – 1200 |
| Vector control coverage | 0 |
| Insecticidal decay half-life | 2 years |
| Bite rate per mosquito per month ($b_m$) | 5-15 |
| Proportion of mosquitoes infected by infectious bite ($p_{mi}$) | 0.37 |
| L3 uptake and development parameter ($\theta_{L3}$) | 4.395 |
| L3 uptake and development parameter ($\sigma_{L3}$) | 0.055 |
| Mosquito death rate per month | 5 |
| Mf birth rate per female worm per month ($\beta_{mf}$) | 1 |
| Proportion L3 leaving mosquito per bite ($\eta_{L3}$) | 0.414 |
| Proportion L3 leaving mosquito that enter host ($\phi_{L3}$) | 0.32 |
| Proportion L3 entering host that develop to adults ($\gamma_{L3}$) | 0.00275 |
| Adult worm death rate per month ($\mu_w$) | 0.0104 |
| Mf death rate per month ($\mu_{mf}$) | 0.1 |
| Host death rate per month ($\mu_h$) | 0.00167 |
| Proportion of mf killed by IA treatment | 0.99 |
| Proportion of adult worms killed by IA treatment | 0.35 |
| Length of worm sterilisation after IA treatment (months) | 9 |
| Reduction in individual bite risk in presence of LLINs (efficacy) | 0.97 |
| Shape parameter for gamma distribution (lymphoedema - $g_l$) | 0.02 |
| Shape parameter for gamma distribution (hydrocele - $g_h$) | 0.71 |

#### *Appendix C.1. Calculation of the prevalence of morbidity (hydrocele, lymphoedema and ADL)*

Each individual is assigned a susceptibility to having hydrocele (if female, this = 0) and lymphoedema. These susceptibilities are drawn from a gamma distribution with a specific shape gl and rate 1/gl (giving the distribution a mean of 1). This susceptibility is drawn at birth and is never changed for the individual. To assess if someone has one of these sequelae, we multiply their susceptibility to the sequelae by the total worms they have had over their lifetime. If this value is greater than a chosen number, then they will be designated as displaying this sequela. A consequence of this is that once a person has a sequela, they will always have it.

For lymphoedema the shape parameter is 0.0033 and for hydrocele it is 0.02 [13]. The number they are compared to is the same for both at 11.3 which was estimated as a fit to a dataset from India [13]. We further assume ADL to occur about twice per year (0–7 times) in 70% (45– 90%) of hydrocele patients, and four (0–7 times) times annually for 95% (90–95%) of patients with lymphoedema [14].

#### *Appendix C.2. Calculation of the DALY averted for morbidity*

In order to estimate the burden of morbidity caused by lymphatic filariasis, we propose the following DALY framework. The burden of the disease can be mathematically estimated as follows,

$$\text{DALY} = \text{YLL} + \text{YLD} \tag{C.1}$$

where YLL is the years lost due to premature death of the disease and YLD is the years lived with disability caused by the disease. We modified this mathematical formula for the estimating the burden of morbidity caused by filariasis as a cause. Consequently, we compute the two components of DALYs as follows:

$$\text{YLD} = \text{Prevalence}_{\text{Morbidity}} \times \text{dw} \tag{C.2}$$

where $\text{Prevalence}_{\text{Morbidity}}$ is the prevalence of morbidity (computed as mentioned in Appendix C.1) and dw is the published disability weights for lymphoedema, hydrocele and ADL. [21]. Now, we consider

$$\text{YLL} = \text{N} \times \text{L} \tag{C.3}$$

where N is the number of deaths caused by the burden of the disease and L is the standard expected life expectancy at the age of death. According to GBD, no deaths occurred as a result of the burden of morbidity caused by lymphatic filariasis. In addition, according to the WHO, although filariasis is one of the leading causes of disability, death from filariasis is rare. Therefore, we will assume N ~ 0. This in turn leads to the years lost due to premature death as zero. Hence, the estimated DALY burden due to morbidity is simply the prevalence of morbidity (computed from Appendix C.1) times the disability weights [26]:

$$\text{DALY} = \text{YLD} = \text{Prevalence}_{\text{Morbidity}} \times \text{dw} \tag{C.4}$$

Therefore, the DALYs averted is computed as,

$$\begin{aligned}\text{DALYs averted} &= \text{DALY burden before starting MDA} \\ &\quad - \text{DALY burden after stopping MDA.}\end{aligned} \tag{C.5}$$

*Appendix C.3. Calculation of the total costs*

The total costs are computed as indicated below,

$$\begin{aligned}\text{Total costs} &= w_1 \times \text{Total MDA rounds} \\ &\quad + w_2 \times \text{Total TAS surevys conducted}\end{aligned} \tag{C.6}$$

where $w_1$ [47] and $w_2$ [7] are the estimated cost weightings computed in accordance with the current purchasing power parity in the US \$. The total MDA rounds and total TAS surveys conducted are computed from the TRANSFIL simulations by keeping a counter for each simulation under the different scenarios. The expected estimated cost weightings of MDA [47] and TAS surveys [7] (excluding the costs for rapid diagnostic tests RDT) for LF programs with annual treatment from the perspective of the endemic country government cover financial and economic costs. The financial costs are the costs of all inputs purchased in cash for MDA, including purchased MDA drugs, materials and supplies, ministry of health personnel salaries, and per diem payments for community drug distributors. Economic costs also include the costs of donated drugs for MDA.

Table C.15: Table with the definitions of the key acronyms used in the manuscript. Adapted from Box 1 in [3].

| Terminologies | Description | Page Defined |
|---|---|---|
| CEA (Cost-Effectiveness Analysis) | A method to compare the relative costs and outcomes (effects) of different interventions or treatments | pg. 1,2,3,11 |
| DALYs (Disability Adjusted Life Years) | The number of years lost due to premature death of the disease and the years lived with disability caused by the disease. | pg. 1,2,3,4,7,8,9,10,11 |
| Disability weights | A factor (between 0 and 1) which is used to calculate the number of years lost due to disability that accounts for the severity of the disease. | pg. 7,8 |
| EINMB (Expected Incremental Net Monetary Benefit) | The mean of the difference in NMB between alternative interventions, a positive incremental NMB indicating that the intervention is cost-effective compared with the alternative at the given willingness-to-pay threshold. In this case, the incremental cost to derive the incremental benefit is less than the maximum amount that the decision-maker would be willing to pay for this benefit. | pg. 3,4,7,9,10,11 |
| EVPI (Expected Value of Perfect Information) | The maximum amount a decision-maker would be willing to pay for perfect information, which would completely eliminate uncertainty. | pg. 4,5,9 |
| EVPPI (Expected Value of Partial Perfect Information) | The value of obtaining information that only partially resolves uncertainty, providing some but not complete clarity. | pg. 4,5,9,10 |
| EVSI (Expected Value of Sample Information) | The value of acquiring information through sampling or additional data collection to reduce uncertainty in decision-making. | pg. 3,5,6,7,9,10 |
| ICER (Incremental Cost-Effective Ratio) | The ratio of change in costs ($\Delta$C) to the change in health impacts ($\Delta$E). In this manuscript, we consider the health impacts due to the DALYs averted for morbidity and the probability of unit increase in elimination. | pg. 1,2,3,4 |
| LOT (Linear Optimal Transport) | A mathematical approach for finding the most cost-effective way to transport resources from one distribution to another while minimizing the total transport cost. Also known as 'Linear Wasserstein Framework' | pg. 1,3,5,6,7,8,9,10,11 |
| LDA (Linear Discriminant Analysis) | A statistical technique (supervised classifier) used for classifying data by identifying linear combinations of features that most effectively distinguish between different classes. | pg. 1,8,9,10 |
| Morbidity | The prevalence or incidence of disease or illness within a population, often measured by the frequency and severity of health-related symptoms. In this study, lymphoedema refers to the improper functioning of the lymph system that results in fluid collection and swelling; hydrocele refers to the swelling of the scrotum due to infection | pg. 1,3,7,8,9,10,11 |
| MDA (Mass Drug Administration) | A method of preventive chemotherapy involving the distribution of anthelminthic drugs to all eligible individuals within a specified area (such as a state, region, province, district, subdistrict, or village) on a routine basis, regardless of their individual infection status. | pg. 2,3,7,8,9,10,11 |
| MDA coverage | The proportion of eligible people in the population who receive MDA. | pg. 3,7,8,9,10,11 |
| NMB (Net Monetary Benefit) | A summary statistic that represents the value of an intervention (in this case, stopping and restarting MDA) in monetary terms when a willingness to pay threshold for a unit of benefit (DALYs averted for morbidity or 1% increase in probability of local elimination) is known. | pg. 4 |
| PCA (Principal Component Analysis) | A dimensionality reduction technique that transforms data into a set of orthogonal components that capture the most variance in the data. | pg. 7,9,10 |
| Probabilistic Sensitivity Analysis (PSA) | A method that examines how uncertainty in the model's input parameters affects the outcomes by running multiple Monte Carlo simulations by varying the input parameters according to their probability distributions. | pg. 2,5 |
| Stopping threshold | A prevalence threshold which is used to determine whether MDA can stop or if further rounds are required. The WHO recommended stopping threshold is <1% mf in children aged 5 years and above or <2% Ag in children aged 6-7 years old. | pg. 1,2,3,7,8,9,10,11 |
| TAS (Transmission Assessment Surveys) | A survey designed to measure whether evaluation units have lowered the prevalence of infection to a level where recrudescence is unlikely to occur, even in the absence of MDA interventions. | pg. 2,3,7,8,10,11 |
| VoI (Value of Information) | The framework used to assess the benefit gained from obtaining additional information to improve decision-making under uncertainty. | pg. 2,3,7 |
| $d_{W^p}(\mu, \nu)$ (Wasserstein distance) | A measure of the difference between two probability distributions, defined as the minimum cost of transporting mass from one distribution to another, where the cost is quantified by the $p$th power of the Euclidean distance function between points in the distributions | pg. 5,6,11 |
| WTP (Willingness To Pay) | The amount of money the decision maker (individual, organization, or government) would be willing to spend per unit of clinical effectiveness (in this case, DALYs averted for morbidity or 1% increase in probability of local elimination). In this study, we rely on the willingness to pay per capita government expenditure for three different example countries | pg. 1,3,4,7,8,9,10,11 |
| Worm burden | The quantity of filarial worms present within a host organism, typically measured by the number or mass of worms within a specific anatomical location or system. | pg. 2,8 |
